# Clinical characteristics of 106 patients with neurological diseases and co-morbid coronavirus disease 2019: a retrospective study

**DOI:** 10.1101/2020.04.29.20085415

**Authors:** Rong Yin, ZhiQi Yang, YaXuan Wei, YuanMing Li, Hui Chen, Zhao Liu, Bo Zhao, DanDan Ma, MeiLing Dan, YingJie Zhang, Xuan Liu, HuiCeng Leng, DaWei Xiang

**Affiliations:** Neurology (Corresponding Author) Neurology Department of The 940th Hospital of Joint Logistic Support Force of the PLA, Lanzhou, China; Huoshenshan Hospital, Wuhan, China; Neurology Department of The 940th Hospital of Joint Logistic Support Force of the PLA, Lanzhou, China; Department of The 940th Hospital of Joint Logistic Support Force of the PLA, Lanzhou, China; Information Department of The 940th Hospital of Joint Logistic Support Force of the PLA, Lanzhou, China; Huoshenshan Hospital, Wuhan, China; Neurology Department of The 940th Hospital of Joint Logistic, Support Force of the PLA, Lanzhou, China; The 940th Hospital of Joint Logistic Support Force of the PLA, Lanzhou, China; Huoshenshan Hospital, Wuhan, China

**Keywords:** neurological disease, COVID-19, Clinical characteristics, mental status

## Abstract

**Objectives:** To describe the clinical characteristics of patients with coronavirus disease 2019 (COVID-19) with co-morbid neurological symptoms.

**Design:** Retrospective case series.

**Setting:** Huoshenshan Hospital in Wuhan, China.

**Participants:** From 4 February to 14 April 2020, 106 patients with neurological diseases were enrolled from all patients in the hospital with confirmed COVID-19 and divided into a severe group and a non-severe group according to their COVID-19 diagnosis.

**Main outcome measures:** Clinical characteristics, laboratory results, imaging findings, and treatment methods were all retrieved through an electronic medical records system and recorded in spreadsheets.

**Results:** The mean (standard deviation, SD) age of patients was 72.7 (11.8) years, and 64 patients were male (60.4%). Among patients with co-morbid neurological diseases, 81 had a previous cerebral infarction (76.4%), 20 had dementia (18.9%), 10 had acute cerebral infarction (9.4%), 5 had sequelae of cerebral haemorrhage (4.7%), 4 had intracranial mass lesions (3.8%), 3 had epilepsy (2.8%), 2 had Parkinson’s disease (1.9%), and 1 had myelopathy (0.9%). Fever (n = 62, 58.5%) was the most common symptom. The most common neurological symptoms were myalgia (n = 26, 24.5%), followed by extremity paralysis (n = 20, 18.9%), impaired consciousness (n = 17, 16%), and positive focal neurological signs (n = 42, 39.6%). Eight patients (7.5%) died. There were more patients with altered mental status in the severe group than in the non-severe group (6 [10.2%] vs. 0, *P* = 0.033). The inflammatory response in the severe group was more significant than that in the non-severe group. There were more patients taking anticoagulant drugs (25 [42.4%] vs. 4 [8.5%], *P* < 0.001) and sedative drugs (22 [37.3%] vs. 9 [19.1%], *P* = 0.041) in the severe group than in the non-severe group. Amid all 93 patients with cerebrovascular diseases, only 32 (34.4%) were taking aspirin, 13 (14%) taking clopidogrel, and 33 (35.5%) taking statins.

**Conclusions:** Patients with COVID-19 with co-morbid neurological diseases had an advanced age, a high rate of severe illness, and a high mortality rate. Among the neurological symptoms, altered mental status was more common in patients with severe COVID-19 with co-morbid neurological diseases.

## Introduction

Coronavirus disease 2019 (COVID-19) due to 2019-novel coronavirus (2019-nCoV) infection has caused a pandemic.^1–3^ From December 2019 to 15 April 2020, the number of COVID-19 patients worldwide reached 2 million, and the mortality has exceeded 120,000.^1^ The number of patients with COVID-19 in most regions of the world is continuing to rise rapidly.^1^ Given the high infectivity of 2019-nCoV, the basic reproduction number of the infection (R0) is between 3.8 and 6.47.^3 4^ The large number of patients with COVID-19 has overwhelmed the health care system, and the mortality rate in some regions exceeds 10%.^1^ Therefore, it is very important to study and understand this unfamiliar disease.

On 11 February 2020, the World Health Organization announced that the disease caused by 2019-nCoV is named COVID-19.^5^ A growing body of evidence has demonstrated that COVID-19 affects not only the respiratory system but also the digestive, urinary, haematological, circulatory, and nervous system.^2,6–16^ Studies have reported neurological symptoms since the onset of COVID-19, including myalgia,^2^ dizziness,^7^ headache,^7^ numbness and weakness in extremities,^13,14^ anosmia and ageusia,^10^ impaired consciousness,^14^ and meningeal irritation,^17^ and these symptoms may even be first-onset, solitary symptoms.^10^ Mao et al.^14^ found that among 214 patients with COVID-19, 78 had neurological symptoms. Clinical case studies of COVID-19 showed that elderly patients and patients with co-morbid neurological diseases had a high rate of severe and critical illness and a high rate of mortality.^2,12,14,15^ Most patients with neurological diseases have co-morbid underlying diseases, including hypertension, diabetes mellitus, coronary heart disease, and metabolic syndrome, which in turn increase their risks of contracting 2019-nCoV, converting into severe illness, and dying.^13,14^ To the best of our knowledge, except for a few case reports, there has been no clinical analysis of patients with neurological diseases and co-morbid COVID-19. Diagnostic information of patients with COVID-19 hospitalised in Huoshenshan Hospital who had completed treatment was retrieved in this study. The clinical characteristics of 106 patients with COVID-19 with co-morbid neurological diseases were analysed.

## Methods

### Study Design and Participants

This study was a single-centre, retrospective, observational study. Huoshenshan Hospital was a temporary special hospital designated by the Chinese government for hospitalising and diagnosing patients with COVID-19. It successively admitted 3,059 patients with confirmed COVID-19 transferred from other hospitals or isolation facilities from 4 February 14 April 2020. Confirmed COVID-19 cases were defined as cases with positive test results of throat swab specimens through high-throughput sequencing or real-time reverse transcription polymerase chain reaction (PCR).^18^ Throat swab specimens were collected, placed in collection tubes containing the virus preservation solution, and then tested by real-time reverse transcription PCR according to the manufacturer’s protocol (Sansure Biotech, Changsha, China). The criteria for clinical classification and cure of COVID-19 were specified in Guidance for Coronavirus Disease 2019 (6th Edition) released by the National Health Commission of Chian, with COVID-19 clinically classified as mild, moderate, severe, or critical.^18^

### Data collection and ethics

Diagnostic information of patients with neurological diseases of acute cerebral infarction, sequelae of cerebral infarction, cerebral haemorrhage, sequelae of cerebral haemorrhage, epilepsy, Parkinson’s disease, Parkinson’s syndrome, dementia, Alzheimer’s disease, peripheral neuropathy, intracranial infection, intracranial mass lesions, cerebral aneurysm, and myelopathy was retrieved by searching the electronic medical records of 3,059 patients admitted to the hospital. After removal of duplicate records, 106 patients were finally enrolled in this study. Through medical records, nursing records, laboratory tests, imaging studies, and medical instructions in the electronic medical records, this study further retrieved the following data and information of the enrolled patients: demographics, history of present illness, symptoms, exposure history, past medical history, history of chronic diseases, neurological symptoms and signs, laboratory test, chest radiographs, chest and head computed tomography, conventional treatment methods (e.g. antiviral treatment, antibiotics, corticosteroids, oxygen support, and symptomatic treatment), special treatment methods (tuzumab, convalescent plasma, stem-cell therapy, and traditional Chinese medicine), prophylactic medication for cerebrovascular diseases, and clinical outcomes. This study also collected the results of laboratory tests and imaging studies first conducted within 48 hours after admission. Upon admission, subjective symptoms were provided by patients who had clear consciousness, normal cognition and psychology, and normal speech; if patients showed impaired consciousness, aphasia, or dementia, the subjective symptoms were provided by their immediate family members who had no problems in cognition, psychology, or speech, and the subjective symptoms were clarified by reviewing previous medical records and communicating directly with their close relatives and physicians. All included data were obtained with verbal consent from the patients or their family members. Each patient’s data were collected and cross-checked by two well-trained and experienced neurologists. In the event of any inconsistency in cross-checking the two sets of data, a third independent expert was designated to review the electronic medical records for data approval. The patients’ data were analysed by a clinical research team comprising epidemiologists. This study was conducted in accordance with the principles of the Helsinki Declaration. This study was approved and written informed consent was waived by the Huoshenshan hospital Ethics Committee on 10 April 2020. owing to enable a rapid response to the novel infectious disease and the urgent need to collect date.

### Statistical analysis

For continuous variables, those conforming to a normal distribution and a non-normal distribution are expressed as mean (standard deviation) and median (interquartile range [IQR]), respectively. Unpaired t-tests and non-parametric tests were performed as appropriate. Categorical variables are expressed as percentages and analysed using the χ^2^ test. Two-sided P-values <0.05 and <0.01 were considered statistically significant and extremely statistically significant, respectively. Data analysis was performed using the SPSS version 19.0 software.

## Results

### Demographic and clinical characteristics of patients with COVID-19 with neurological diseases

A total of 3,059 patients with confirmed COVID-19 were hospitalised in Huoshenshan Hospital, including 1,417 (46.3%) severe cases and 149 (4.9%) critically ill cases, and 69 (2.3%) subsequently died. A total of 106 hospitalised patients with COVID-19 and co-morbid neurological diseases were enrolled for analysis. Their demographics, clinical characteristics, and outcomes are shown in Table 1. Their average age was 72.7 (11.8) years, 86 patients (81.1%) were older than 65 years, and 64 patients (60.4%) were male. The number of patients with co-morbid hypertension, diabetes mellitus, coronary heart disease, and chronic obstructive pulmonary disease were 72 (67.9%), 37 (34.9%), 28 (26.4%), and 14 (13.2%), respectively. Amid the neurological co-morbidities, a previous cerebral infarction was most common, occurring in 81 patients (76.4%); dementia occurred in 20 patients (18.9%), acute cerebral infarction in 10 (9.4%), sequelae of cerebral haemorrhage in 5 (4.7%), intracranial mass lesions in 4 (3.8%), epilepsy in 3 (2.8%), Parkinson’s disease in 2 (1.9%), and myelopathy in 1 (0.9%). Fever (n = 62, 58.5%) was still the most common symptom. Other common non-neurological symptoms were as follows: coughs in 61 patients (57.5%), fatigue in 53 patients (50%), chest tightness in 42 patients (39.6%), expectoration in 34 patients (32.1%), diarrhoea in 17 patients (16%), and dyspnoea in 12 patients (11.3%). The most common neurological symptoms were myalgia in 26 patients (24.5%), extremity paralysis in 20 patients (18.9%), and impaired consciousness in 17 patients (16%). Positive focal neurological signs occurred in 42 patients (39.6%). Headache (n = 8, 7.5%), nausea (n = 6, 5.7%), vomiting (n = 7, 6.6%), dizziness (n = 9, 8.5%), altered mental state (n = 6, 5.7%), and exacerbated neurological symptoms (n = 8, 7.5%) all occurred in this enrolled group. The mean pulse oxygen saturation was 91% (6%). The mean duration from onset to cure was 37 (17) days. Eight patients (7.5%) died.

**Table 1.** Clinical characteristics of patients with COVID-19 with co-morbid neurological diseases.

| | Total ($n = 106$) | Non-severe ($n = 47$) | Severe ($n = 59$) | $P$ value |
| --- | --- | --- | --- | --- |
| **Age, median (SD), years** | 72.7 (11.8) | 71.9 (12.1) | 73.5 (11.5) | 0.471 |
| <65 years | 20 (18.9) | 11 (23.4) | 9 (15.3) | 0.287 |
| ≥65 years | 86 (81.1) | 36 (76.6) | 50 (84.7) |  |
| **Sex, n (%)** |  |  |  | 0.517 |
| Female | 42 (39.6) | 17 (36.2) | 25 (42.4) |  |
| Male | 64 (60.4) | 30 (63.8) | 34 (57.6) |  |
| **Close contact with confirmed case history, n (%)** | 98 (92.5) | 45 (95.7) | 53 (90) | 0.252 |
| **Smoking history, n (%)** | 18 (17.0) | 6 (12.8) | 12 (20.3) | 0.302 |
| **Coexisting disorder, n (%)** |  |  |  |  |
| Hypertension | 72 (67.9) | 33 (70.2) | 39 (66.1) | 0.652 |
| Diabetes | 37 (34.9) | 18 (38.3) | 19 (32.2) | 0.513 |
| Hyperlipemia | 2 (1.9) | 1 (2.1) | 1 (3.4) | 1 |
| Pulmonary heart disease | 3 (2.8) | 2 (4.3) | 1 (1.7) | 0.841 |
| Coronary artery disease | 28 (26.4) | 14 (29.8) | 14 (23.7) | 0.482 |
| Atrial fibrillation | 6 (5.7) | 3 (6.4) | 3 (5.1) | 1 |
| Tuberculosis | 5 (4.7) | 3 (6.4) | 2 (3.4) | 0.794 |
| Chronic bronchitis | 6 (5.7) | 1 (2.1) | 5 (8.5) | 0.326 |
| COPD | 14 (13.2) | 6 (12.8) | 8 (13.6) | 0.905 |
| Cancer<sup>b</sup> | 9 (8.5) | 1 (2.1) | 8 (13.6) | 0.081 |
| Chronic gastritis | 8 (7.5) | 5 (10.6) | 3 (5.1) | 0.481 |
| Chronic liver disease | 6 (5.7) | 3 (6.4) | 3 (5.1) | 1 |
| Chronic kidney disease | 6 (5.7) | 2 (4.3) | 4 (6.8) | 0.892 |
| Metabolic disease | 8 (7.5) | 2 (4.3) | 6 (10.2) | 0.438 |
| **Neurological co-morbidities, n (%)** |  |  |  |  |
| Acute ischemic stroke | 10 (9.4) | 4 (8.5) | 6 (10.2) | 1 |
| Remote cerebral infarction | 81 (76.4) | 35 (74.5) | 46 (78) | 0.673 |
| Haemorrhage sequelae | 5 (4.7) | 2 (4.3) | 3 (5.1) | 1 |
| Dementia | 20 (18.9) | 7 (14.9) | 13 (22) | 0.351 |
| Intracranial mass lesion | 4 (3.8) | 1 (2.1) | 3 (5.1) | 0.779 |
| Parkinson | 2 (1.9) | 0 (0) | 2 (3.4) | NA |
| Epilepsy | 3 (2.8) | 0 (0) | 3 (5.1) | NA |
| Myelenterosis | 1 (0.9) | 1 (2.1) | 0 (0) | NA |
| **Signs and symptoms, n (%)** |  |  |  |  |
| Fever | 62 (58.5) | 25 (53.2) | 37 (62.7) | 0.323 |
| Cough | 61 (57.5) | 25 (53.2) | 36 (61.0) | 0.418 |
| Chest distress | 42 (39.6) | 16 (34.0) | 26 (44.1) | 0.294 |
| Weak | 53 (50.0) | 22 (46.8) | 31 (52.5) | 0.558 |
| Expectoration | 34 (32.1) | 8 (17.0) | 26 (44.1) | 0.003 |
| Dyspnoea | 12 (11.3) | 0 (0) | 12 (20.3) | 0.001 |
| Diarrhoea | 17 (16.0) | 4 (8.5) | 13 (22.0) | 0.59 |
| ARDS | 3 (2.8) | 0 (0) | 3 (5.1) | NA |
| Shock | 5 (4.7) | 1 (2.1) | 4 (6.8) | 0.508 |
| Acute renal failure | 7 (6.6) | 1 (2.1) | 6 (10.2) | 0.207 |
| Acute cardiac injury | 7 (6.6) | 1 (2.1) | 6 (10.2) | 0.207 |
| Acute liver lesion | 8 (7.5) | 2 (4.2) | 6 (10.2) | 0.438 |
| **Nervous system symptoms, n (%)** |  |  |  |  |
| Myalgia | 26 (24.5) | 13 (27.7) | 13 (22.0) | 0.504 |
| Headache | 8 (7.5) | 1 (2.1) | 7 (11.9) | 0.13 |
| Nausea | 6 (5.7) | 2 (4.3) | 4 (6.8) | 0.892 |
| Vomit | 7 (6.6) | 2 (4.3) | 5 (8.5) | 0.635 |
| Dizziness | 9 (8.5) | 4 (8.5) | 5 (8.5) | 1 |
| Hemiplegia | 20 (18.9) | 7 (14.9) | 13 (22.0) | 0.351 |
| Insanity | 6 (5.7) | 0 (0) | 6 (10.2) | 0.033 |
| Progressive neurologic symptom | 8 (7.5) | 1 (2.1) | 7 (11.9) | 0.13 |
| Disturbance of consciousness | 17 (16.0) | 4 (8.5) | 13 (22.0) | 0.059 |
| Positive signs of nervous system | 42 (39.6) | 16 (34.0) | 26 (44.1) | 0.294 |
| **Vital signs on admission, median (SD)** |  |  |  |  |
| Respiratory rate | 20 (2) | 20 (2) | 21 (3) | 0.195 |
| Pulse rate | 88 (14) | 86 (13) | 89 (15) | 0.375 |
| SaO<sub>2</sub>, % | 91 (6) | 95 (1) | 88 (7) | <0.01 |
| SBP | 134 (19) | 136 (17) | 132 (21) | 0.257 |
| DBP | 80 (15) | 82 (13) | 78 (17) | 0.049 |
| **Time of therapy (days), median (SD)** | 37 (17) | 37 (16) | 38 (18) | 0.963 |
| **Clinical outcome, n (%)** |  |  |  | 0.008 |
| Cure | 98 (92.5) | 47 (100) | 51 (86.4) |  |
| Death | 8 (7.5) | 0 (0) | 8 (13.6) |  |

<sup>a</sup>*P* values indicate differences between patients with severe and non-severe disease. *P* < 0.05 was considered statistically significant.
<sup>b</sup>Included in this category is any type of cancer.
ARDS = acute respiratory distress syndrome; COPD, chronic obstructive pulmonary disease; DBP, diastolic blood pressure; NA = not applicable; SaO<sub>2</sub> = oxygen saturation of arterial blood; SBP, systolic blood pressure.

Disease was classified clinically as mild (n = 0), moderate (n = 47), severe (n = 48), and critical illness (n = 11). The patients with mild and moderate disease were combined into a non-severe group, totalling 47 patients (44.3%). Patients with severe disease and critically ill patients were combined into the severe group, totalling 59 patients (55.7%). Comparisons were conducted between the two groups (Table 1). There was no significant difference in age between the severe group and the non-severe group (73.5 (11.5) vs. 71.9 (12.1) years, *P* = 0.47). There was a higher percentage of male vs. female patients in both groups: 57.6% (n = 34) vs. 63.8% (n = 30), respectively (*P* = 0.517). In terms of past medical history, there was no significant difference in either neurological diseases or other underlying diseases between the two groups. The severe group had more cases of dyspnoea (12 (20.3%) vs. 0, *P* = 0.001) and expectoration (26 (44.1%) vs. 8 (17%), *P* = 0.003) than the non-severe group. Per neurological symptoms, the severe group had significantly more patients with altered mental state than the non-severe group (6 (10.2%) vs. 0, *P* = 0.033), and there were no significant differences in other neurological symptoms and general symptoms between the two groups. With respect to vital signs after hospitalisation, the pulse oxygen saturation in the severe group was 88% (7%), significantly lower than 95% (1%) in the non-severe group (*P* < 0.01). The systolic and diastolic blood pressure values in the severe group (132 (21) and 78 (17)) were lower than those in the non-severe group (136 (17) and 82 (13)), and the difference in diastolic pressure between the two groups was statistically significant (*P* = 0.049). All 8 patients (13.6%) who subsequently died were in the severe group, and the percentage was significantly higher than that in the non-severe group *(P* = 0.008). Amid the 8 deceased patients, 7 were critically ill before death (7/11; 63.64%), and 1 was severely ill (1/48; 2.08%).

### Laboratory and imaging findings of patients with COVID-19 with co-morbid neurological diseases

Table 2 presents the laboratory and imaging findings. The inflammatory response in the severe group was more significant than that in the non-severe group, including a decrease in red blood cell count (3.51 [IQR, 3.04-3.95] × 10^12^/L vs. 4.09 [IQR, 3.46-4.42] × 10^12^/L, *P* = 0.001), an increase in percentage of neutrophils (73.4% [IQR, 63.6%-81.5%] vs. 63.6% [IQR, 59.6%-68%], *P* < 0.001), a decrease in percentage of lymphocytes (16.2% [IQR, 10.3%-24.2%] vs. 23.9 [IQR, 19.1%-26.9%], *P* = 0.001), and a decrease in percentage of monocytes (6.8% [IQR, 4.4%-8.6%] vs. 8.1% [IQR, 6.6%-10%], *P* = 0.029). The percentage of patients with significantly high levels of C-reactive protein (CRP) (81.4% (48/59] vs. 39.1% (18/46], *P* < 0.001), hypersensitive CRP (66.1% (39/59] vs. 19.6% (9/46], *P* = 0.011), procalcitonin (69% (41/54] vs. 48.5% (16/33], *P* = 0.047), and interleukin-6 (62.5% (26/40] vs. 37.5% (9/24], *P* = 0.032) was higher in the severe group than in the non-severe group. The albumin level of the entire study population was lower than normal (35.85 [IQR, 32.53-39.23] g/L), and the value in the severe group was lower than that in the non-severe group (33.6 [IQR, 30.2-36.8] g/L vs. 37.4 [IQR, 34.8-41.1] g/L, *P* < 0.001). The values of other biochemical indicators, including alanine aminotransferase, aspartate aminotransferase, blood glucose, creatinine, urea nitrogen, uric acid, lactate dehydrogenase, creatine kinase, creatine kinase isoenzyme, and serum potassium, sodium, chloride, and calcium in either the entire study population or the severe and non-severe groups were all in the normal reference range. Although there were statistically significant differences in the levels of aspartate aminotransferase, uric acid, lactate dehydrogenase, and serum calcium between the severe and non-severe groups, there was no clinical significance because the levels were within the clinical normal reference range. In terms of blood coagulation, the D-dimer level increased in the entire study population (0.95 [IQR, 0.52-1.835] mg/L), with a significantly higher level in the severe group than in the non-severe group (1.05 [IQR, 0.69-2.36] mg/L vs. 0.69 [IQR, 0.38-1.33] mg/L). The prothrombin time and activated partial thromboplastin time in the entire study population, severe group, and non-severe group were all in the normal reference range. The prothrombin time in the severe group was longer than that in the non-severe group (13.58 [IQR, 12.54-15.02 s] vs. 13.02 [IQR, 12.03-13.76] s, *P* = 0.011). Chest imaging findings showed no difference in the upper and lower lobes between the two groups in terms of locations of lung infections. However, the incidence of pleural effusion in the severe group was higher than that in the non-severe group (21 (35.6%] vs. 4 (8.5%], *P* = 0.001).

**Table 2.**
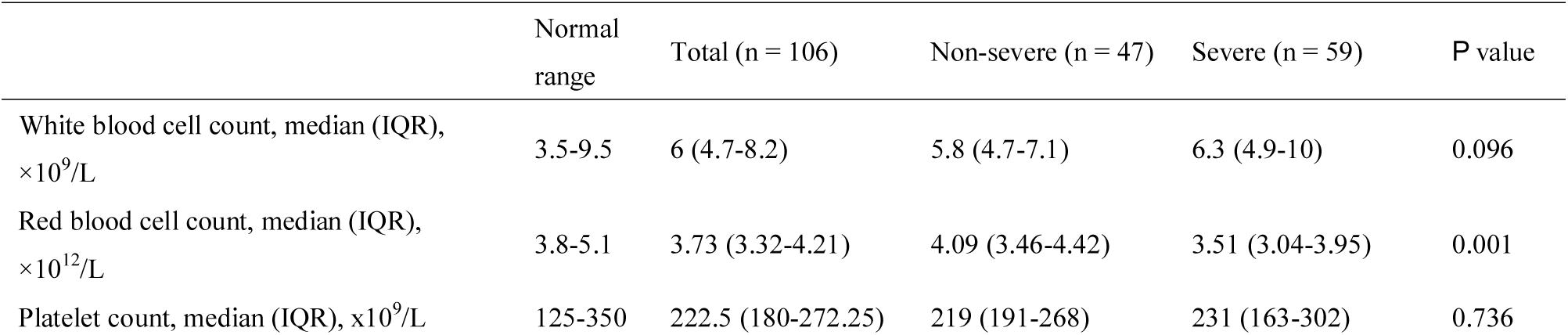
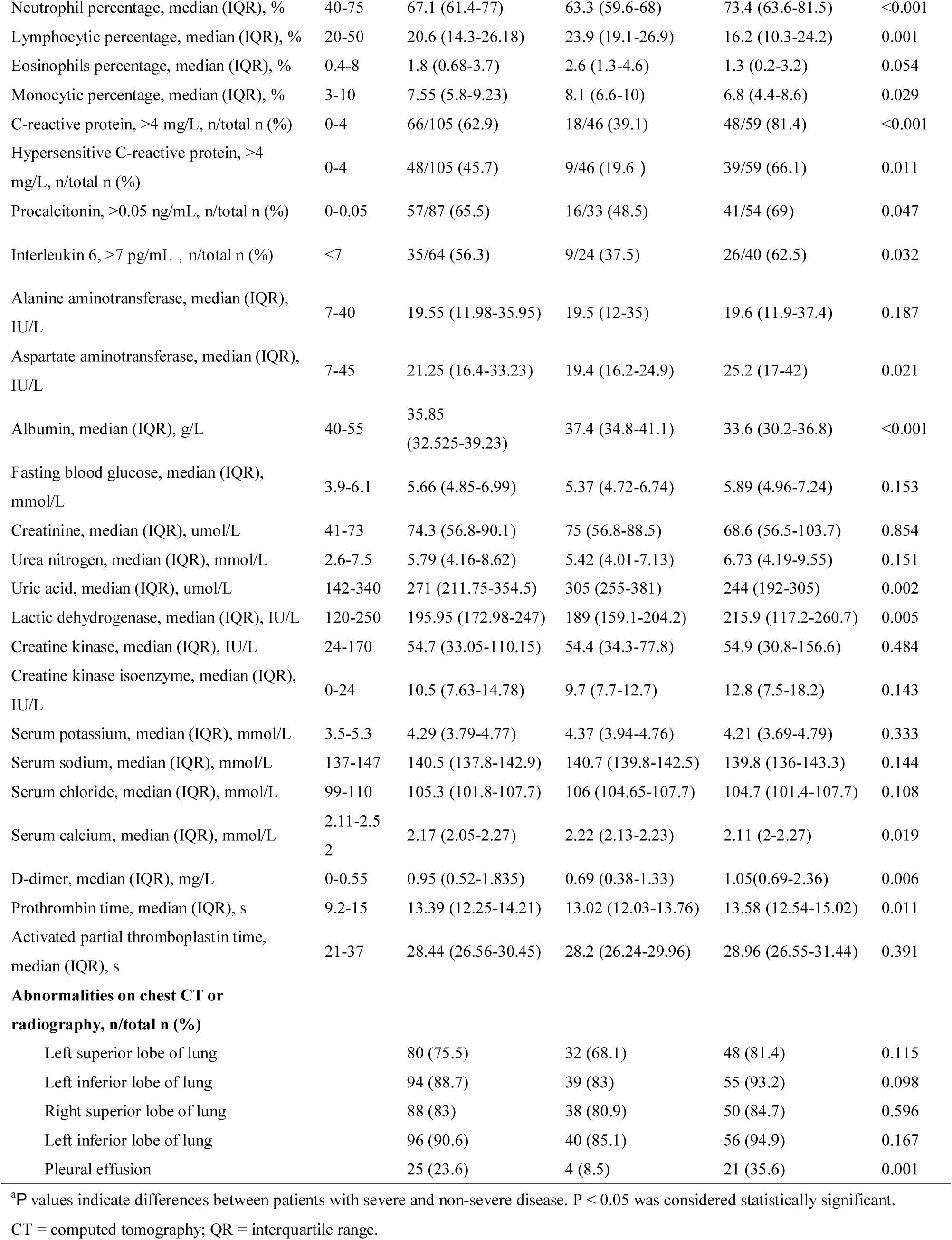
Laboratory and imaging findings of patients with COVID-19 with co-morbid neurological diseases.

### Treatment of patients with COVID-19 with co-morbid neurological diseases

Table 3 shows that 87 out of 106 patients (82.1%) were administered nasal catheter oxygenation, 14 (13.2%) were administered face mask oxygenation, 15 (14.2%) were administered high-flow nasal catheter oxygenation, 13 (12.3 %) were treated using a non-invasive ventilator, and 9 (8.5%) required mechanical ventilation with endotracheal intubation. The proportion of patients administered face mask oxygenation, high-flow nasal catheter oxygenation, and non-invasive and invasive mechanical ventilation was significantly higher in the severe group than in the non-severe group. Only 1 patient (2.1%) in the non-severe group was administered face mask oxygenation and high-flow humidified oxygenation, and no patients were treated using a non-invasive ventilator or required mechanical ventilation. In the severe group, the number of patients administered face mask oxygenation, high-flow nasal catheter oxygenation, and non-invasive and invasive mechanical ventilation were 13 (22%), 14 (23.7%), 13 (22%), and 9 (15.2%), respectively. Regarding antiviral treatment, there was no significant difference in the use of arbidol and interferon inhalation between the two groups. The proportion of patients taking antibiotics was significantly higher in the severe group than in the non-severe group (moxifloxacin: 26 [44.1%] vs. 13 [27.7%], *P* = 0.082; cefoperazone-sulbactam sodium: 16 [27.1%] vs. 2 [4.3%], *P* = 0.002; meropenem: 10 [16.9%] vs. 1 [2.1%], *P* = 0.03). The proportion of patients taking expectorants and steroids was higher in the severe group than in the non-severe group (expectorants: 22 [37.3%] vs. 7 (14.9%], *P* = 0.01; steroids: 18 [30.5%] vs. 4 [8.5%], *P* = 0.006). In the severe group, 8 patients (13.6%) were on intravenous gamma immunoglobulin, 9 patients (15.3%) used convalescent plasma, 2 patients (3.4%) received stem cell therapy, and 3 patients (5.1%) received continuous renal replacement therapy. Among the entire study population, 77 patients (72.6%) were on traditional Chinese medicine, 29 (27.4%) on anticoagulant drugs, and 31 (29.2%) on sedative drugs. The severe group had a higher percentage of patients taking anticoagulant drugs than the non-severe group (25 [42.4%] vs. 4 [8.5%], *P* < 0.001) and sedative drugs (22 [37.3%] vs. 9 [19.1%], *P* = 0.041),. In this study, 93 patients had a cerebrovascular disease. Regarding prophylactic medication for stroke, only 32 of 93 patients (34.4%) were on aspirin, 13 of 93 (14%) on clopidogrel, and 33 of 93 (35.5%) on statins.

**Table 3.** Treatments for patients with COVID-19 with co-morbid neurological diseases.

|  | Total (n = 106) | Non-severe (n = 47) | Severe (n = 59) | *P* value<sup>a</sup> |
| --- | --- | --- | --- | --- |
| **Oxygen therapy, n (%)** |  |  |  |  |
| Nasal cannula | 87 (82.1) | 38 (80.9) | 49 (83) | 0.769 |
| Mask inhalation of oxygen | 14 (13.2) | 1 (2.1) | 13 (22) | 0.003 |
| High-flow nasal cannula | 15 (14.2) | 1 (2.1) | 14 (23.7) | 0.002 |
| Non-invasive mechanical ventilation | 13 (12.3) | 0 (0) | 13 (22) | 0.001 |
| Invasive mechanical ventilation | 9 (8.5) | 0 (0) | 9 (15.2) | 0.014 |
| **Glucocorticoid therapy, n (%)** | 22 (20.8) | 4 (8.5) | 18 (30.5) | 0.006 |
| **Antibiotics, n (%)** |  |  |  |  |
| Moxifloxacin | 39 (36.8) | 13 (27.7) | 26 (44.1) | 0.082 |
| Cefperazone-sulbactam | 18 (17) | 2 (4.3) | 16 (27.1) | 0.002 |
| Meropenem | 11 (10.4) | 1 (2.1) | 10 (16.9) | 0.03 |
| **Antiviral therapy, n (%)** |  |  |  |  |
| Arbidol hydrochloride | 50 (47.2) | 21 (44.7) | 29 (49.2) | 0.467 |
| Interferon atomisation | 6 (5.7) | 3 (6.4) | 3 (5.1) | 1 |
| **Special treatment, n (%)** |  |  |  |  |
| Tocilizumab | 6 (5.7) | 2 (4.3) | 4 (6.8) | 0.892 |
| Convalescent plasma therapy | 9 (8.5) | 0 (0) | 9 (15.3) | 0.014 |
| Stem cell treatment | 2 (1.9) | 0 (0) | 2 (3.4) | 0.502 |
| **Supportive treatment, n (%)** |  |  |  |  |
| Antitussive | 33 (31.1) | 12 (25.5) | 21 (35.6) | 0.266 |
| Expectorant | 29 (27.4) | 7 (14.9) | 22 (37.3) | 0.01 |
| Thymosin | 28 (26.4) | 9 (19.1) | 19 (32.2) | 0.13 |
| Intravenous immunoglobulin | 8 (7.5) | 0 (0) | 8 (13.6) | 0.024 |
| **Continuous renal replacement therapy, n (%)** | 3 (2.8) | 0 (0) | 3 (5.1) | 0.328 |
| **Extracorporeal membrane oxygenation, n (%)** | 0 (0) | 0 (0) | 0 (0) | NA |
| **Traditional Chinese medicine, n (%)** | 77 (72.6) | 35 (74.5) | 42 (71.2) | 0.707 |
| **Central nervous system drugs, n/total n (%)** |  |  |  |  |
| Aspirin | 32/93 (34.4) | 10/40 (25) | 22/53 (41.5) | 0.097 |
| Clopidogrel | 13/93 (14) | 7/40 (17.5) | 6/53 (11.3) | 0.395 |
| Statins | 33/93 (35.5) | 14/40 (35) | 19/53 (35.8) | 0.932 |
| Anticoagulant therapy | 29/106 (27.4) | 4/47 (8.5) | 25/59 (42.4) | <0.001 |
| Sedative agents | 31/106 (29.2) | 9/47 (19.1) | 22/59 (37.3) | 0.041 |

<sup>a</sup>*P* values indicate differences between patients with severe and non-severe disease. *P* < 0.05 was considered statistically significant.

## Discussion

This study presented an analysis of the clinical characteristics of neurological diseases co-morbid with COVID-19 in the largest sample size so far. Of the 106 patients enrolled in this study, 59 (55.7%) had severe and 47 (44.3%) had non-severe disease, and the average age was 72.7 (11.8) years. Fever was still the most common non-neurological symptom. However, the proportion of patients with fever was only 58.5%, which was significantly lower than the 85.14%-98.6% reported by other studies,^2 12 15 16 19^ but similar to the proportion of 61.7% in 214 patients in a study by Mao et al.^14^ Mo et al.^19^ also found that the proportion of patients with fever in the refractory group was 74.1%, lower than the 90% in the moderate group, and the average age of the refractory group was 61 years, significantly higher than the 46 years of the moderate group. The low occurrence of fever in this group was linked to an advanced average age, a decrease in pulmonary defence function, and a decline in systemic immunity. In this study, the proportions of patients with expectoration and dyspnoea in the severe group were significantly higher than those in the non-severe group, and moreover, the severe group had significantly higher neutrophil ratios and higher levels of infection indicators (CRP, hypersensitive CRP, and procalcitonin). This suggested that the severe group was likely to have a higher rate of co-morbid bacterial infections than the non-severe group, which in turn explained why there was a higher proportion of patients using mechanical ventilation, non-invasive ventilators, high-flow humidified oxygenation, face mask oxygenation, expectorants, antibiotics, and steroids in the severe group than in the non-severe group. Owing to co-morbid neurological diseases, most patients had different degrees of motor dysfunction, resulting in long-term bed rest and fluid accumulation in the dorsal regions of the lungs, which was more likely to cause hypostatic pneumonia and reduce pulmonary reserve. In the meantime, bulbar palsy further exacerbated the inflammation of the lungs. In addition to the fact that patients had many co-morbidities at an advanced age, the above observations could be a main reason why there was a higher mortality rate (13.6%) in patients with severe COVID-19 with co-morbid neurological diseases than in other populations.^2 7 19^

In this study, altered mental status occurred in 6 patients (5.7%), all of whom were in the severe group. Our research team has reported a case of COVID-19 presenting with intracranial infection that resulted in mental and behavioural abnormalities.^17^ Helms et al.^20^ performed head magnetic resonance imaging (MRI) on 13 patients with severe COVID-19 and found that 8 patients exhibited cerebral leptomeningeal enhancement, and the electroencephalogram (EEG) of 1 patient showed diffuse slow waves in the bilateral frontal lobes, which was similar to the EEG of encephalopathy. Although no patient presented with meningeal irritation and obvious inflammatory changes in the cerebrospinal fluid, the above observation was still highly indicative of meningeal involvement. This suggested that 2019-nCoV could involve the central nervous system (CNS), especially in patients who had blood-brain barrier breakdown due to a neurological disease. The pathophysiology pathways of 2019-nCoV involvement in the nervous system is still unclear. The viruses 2019-nCoV, SARS-CoV, and MERS-CoV are all coronaviruses, and they have highly homologous sequences. Most coronaviruses have similar viral structures and transmission routes.^21^ A previous study revealed that SARS-CoV could invade the mouse brain tissue through olfactory epithelial cells in the nasal cavity, resulting in neuronal death.^22^ Autopsy results also revealed that SARS-CoV existed in the CNS.^22^ With the in-depth understanding of COVID-19, it has been reported that patients with COVID-19 could experience olfactory and taste impairment.^10^ Therefore, 2019-nCoV invasion of the brain tissue through the lamina cribrosa close to the olfactory bulb may be a pathway of CNS infection.^23^ Olfactory and taste impairment was not observed in the 106 participants in the present study, suggesting that the above-mentioned pathway may not be the main pathway for the virus invasion of the CNS. Another possible reason for the lack of olfactory and taste impairment may be that there were few frontline neurologists and thus such symptoms were likely to be omitted.

Angiotensin-converting enzyme 2 (ACE2) has been identified as a functional receptor of SARS-CoV.^24^ It was reported that ACE2 receptors are expressed on glial cells and neurons in the brain.^23^ Hamming et al.^25^ studied ACE2 protein localisation in different organs of the human body and found that ACE2 existed in the endothelial cells of arteries and veins as well as arterial smooth muscle cells of all organs. It is difficult for the virus to invade the CNS owing to the presence of the blood-brain barrier. However, the meninges are rich in blood vessels, which may explain why cerebral leptomeningeal enhancement appeared in 8 patients with severe COVID-19 in a previous report. ACE2 receptors also appear in skeletal muscles. It was found in this study that 26 patients (24.5%) had myalgia, with no significant difference between the two groups and with lactate dehydrogenase levels in the normal range. However, the lactate dehydrogenase level was significantly higher in the severe group than in the non-severe group, which further confirmed that this kind of injury may be related to ACE2 in skeletal muscles. This result was similar to that of Mao et al.,^14^ but it still remains to be verified by conducting autopsies to find corresponding evidence in skeletal muscles. Meanwhile, it cannot be ruled out that infection may cause an excessive immune response, damaging the nervous system, and an increase in the level of cytokines may damage skeletal muscles.^24^

Recent reports have shown that 2019-nCoV infection may cause a cytokine storm, resulting in multiple organ dysfunction, including cellular immunity deficiencies, coagulation activation, myocardial injury, liver injury, and kidney injury.^11,14,15^ In the present study, the severe group had a decrease in red blood cells, lymphocytes, and monocytes as well as an increase in interleukin-6 levels, prothrombin time, and D-dimer levels. This suggested that the patients with severe disease were more likely to develop a cytokine storm, which caused coagulation activation and an increase in red blood cell destruction. The elevated D-dimer levels also increase the risk of cerebrovascular disease.^26^ Although the proportion of patients taking anticoagulant drugs in the severe group was significantly higher than that in the non-severe group, the proportion of patients on prophylactic medications (anti-platelets and statins) for secondary prevention of cerebrovascular disease was very low in the 106 patients, which may be a reason for the high incidence of acute cerebrovascular disease (9.4%) and high mortality rate of patients with COVID-19 with co-morbid neurological diseases in this study. The proportion of patients taking sedative drugs was high in the severe group, mainly because patients needed to be sedated when undergoing mechanical ventilation. After these data were excluded, there was no significant difference between the two groups. However, it should be noted that patients with neurological diseases may develop central respiratory depression, which is also a cause of increasing mortality.^20,23^

## Limitations

This study had several limitations. First, it was a retrospective study, with a sample size of only 106 cases, which may lead to a bias in clinical observation. Second, given that some patients had dementia, aphasia, and impaired consciousness, it was inevitable that some information failed to be collected. In an infectious disease hospital, the collection of neurological symptoms may be inaccurate and incomplete, and when extracting data from electronic medical records, clinical symptoms may be underestimated. For example, smell and taste impairment and peripheral nerve damage were not observed in this study. Third, because there was no MRI equipment in a temporarily built hospital, it was impossible to verify the affected areas in the CNS, and to avoid the risk of infection, a lumbar puncture for cerebrospinal fluid analysis was rarely performed, and the diagnosis rate of intracranial infection was low. Furthermore, owing to a lack of neuroelectrophysiological testing, it was impossible to determine peripheral nerve injury. Fourth, the completion rate of a blood gas analysis was low in the non-severe group, and this index was removed to avoid clinical bias. Finally, considering that there were only 11 critically ill patients, their clinical characteristics were not compared with those of other patients, as the comparison would otherwise introduce inter-group bias, but this practice may prevent a deeper insight into the clinical data.

## Conclusions

In summary, patients with COVID-19 with co-morbid neurological diseases had an advanced age, a high rate of severe illness, and a high mortality rate. Regarding neurological symptoms, altered mental state was more common in patients with severe COVID-19 with co-morbid neurological diseases. For patients with neurological diseases who have no typical respiratory symptoms, it is necessary to pay attention to neurological symptoms and the physical examination, improve nucleic acid testing for virus in a timely manner, and perform pulmonary imaging studies, which would be beneficial in the early detection of COVID-19 and avoidance of infection. Moreover, attention should be paid to the prevention and treatment of primary neurological diseases, which would help to reduce the mortality of such patients.

### What is already known on this topic

As of 15April, more than 2 million patients had been infected with COVID-19 worldwide, and the death toll exceeded 120,000.

COVID-19 can involve the nervous system and affect the outcome.

The clinical characteristics of COVID-19 co-morbid with neurological diseases have not yet been reported in a large sample size.

### What this study adds

Patients with COVID-19 with co-morbid neurological diseases had an advanced age, a high rate of severe illness, and a high mortality rate.

It is necessary to pay attention to patients with COVID-19 with altered mental states.

During the diagnosis and treatment of patients with COVID-19, it is necessary to pay attention to the history and physical exam of the nervous system as well as the prevention and treatment of primary neurological diseases.

We thank all the patients and their families involved in this study, as well as many doctors, nurses, and civilians working together to fight against COVID-19.

## Data Availability

The data used to support the findings of this study are available from the corresponding author upon request.

## Contributors

RY, ZQY, YXW, YML, HC, ZL and BZ contributed equally to this paper, as did RY, ZQY, HC and DWX designed the study, had full access to all data in the study, and takes responsibility for the integrity and accuracy of the data analysis. RY, ZQY, HC, DDM, MLD, YJZ, XL and HCL contributed to patient recruitment, data collection, data analysis, data interpretation, literature search, and writing of the manuscript. DDM, MLD, YJZ, XL and HCL had roles in patient recruitment, data collection, and clinical management. RY, ZL, BZ, DDM, MLD, YJZ, XL and HCL had roles in the patient management, data collection, data analysis, and data interpretation. All authors contributed to data acquisition, data analysis, or data interpretation, and all reviewed and approved the final version of the manuscript. The corresponding author attests that all listed authors meet authorship criteria and that no others meeting the criteria have been omitted. RY is the guarantor.

## Ethical approval

The case series was approved by the Institutional Review Board of The 940th Hospital of Joint Logistic Support Force of the PLA (2020KYLL080). Written informed consent was waived owing to the rapid emergence of this infectious disease.

## Data sharing

No additional data available.

## Transparency declaration

The lead author (the manuscript’s guarantor) affirms that the manuscript is an honest, accurate, and transparent account of the study being reported; that no important aspects of the study have been omitted; and that any discrepancies from the study as planned (and, if relevant, registered) have been explained.

## Dissemination to participants and related patient and public communities

No study participants were involved in the preparation of this article. The results of the article will be presented at relevant conferences.

